# Enhanced Gut Microbiome Capacity for Amino Acid Metabolism is associated with Peanut Oral Immunotherapy Failure

**DOI:** 10.1101/2024.07.15.24309840

**Authors:** Mustafa Özçam, Din L. Lin, Chhedi L. Gupta, Allison Li, Lisa M. Wheatley, Carolyn H. Baloh, Srinath Sanda, Stacie M. Jones, Susan V. Lynch

## Abstract

Peanut Oral Immunotherapy (POIT) holds promise for remission of peanut allergy, though treatment is protracted and successful in only a subset of patients. Because the gut microbiome is linked to food allergy, we sought to identify fecal microbial predictors of POIT efficacy and to develop mechanistic insights into treatment response. Longitudinal functional analysis of the fecal microbiome of children (n=79) undergoing POIT in a first double-blind, placebo-controlled clinical trial, identified five microbial-derived bile acids enriched in fecal samples prior to POIT initiation that predicted treatment efficacy (AUC 0.71). Failure to induce disease remission was associated with a distinct fecal microbiome with enhanced capacity for bile acid deconjugation, amino acid metabolism, and increased peanut peptide degradation *in vitro*. Thus, microbiome mechanisms of POIT failure appear to include depletion of immunomodulatory secondary bile and amino acids and the antigenic peanut peptides necessary to promote peanut allergy desensitization and remission.

---

Peanut protein allergy (PA) is a life-threatening condition affecting 2% of the population in industrialized nations^1^. A leading cause of food-induced anaphylaxis^2^, the condition was not treatable until the approval of peanut oral immunotherapy (Palforzia^TM^) by the U.S. Food and Drug Administration in 2020, with strict avoidance of peanuts and peanut-containing products representing the most effective disease management strategy^3^. Peanut oral immunotherapy (POIT) has emerged as a treatment that is widely utilized for PA^4^. Gradual oral introduction of increasing concentrations of peanut product induces desensitization, defined as an increase in reaction threshold while on treatment, in approximately 50-70% of treated patients. While POIT has demonstrated efficacy in desensitizing patients to peanut, the induction of remission, defined as the prolonged absence of clinical reactivity after treatment cessation, is observed in a smaller subset of approximately 20-30% of POIT-treated patients^4–6^. The cost, prolonged duration (years), and burden of daily treatment associated with POIT highlight the need for improved predictive biomarkers and adjunctive treatments to increase efficacy.

The IMPACT, Oral Immunotherapy for the Induction of Tolerance and Desensitization in Peanut-Allergic Children trial (NCT01867671) was the first randomized, double-blinded, placebo-controlled, multicenter clinical trial to evaluate the efficacy and safety of POIT in peanut-allergic children ages 12-48 months old. While 71% of the children receiving POIT achieved desensitization, only 21% of children achieved remission following POIT discontinuation and 26-weeks of peanut avoidance. The IMPACT trial yielded three clinical outcomes among peanut-allergic children treated with POIT: **i)** those who achieved both desensitization and remission (D+R+), **ii)** those who achieved only desensitization but not remission (D+R-), **iii)** those who did not achieve desensitization or remission (D-R-). Notably, younger age and lower peanut-specific serum IgE concentrations at the outset of the trial were more likely to result in a D+R+ outcome^7^. Both age^8^ and allergic sensitization status^9^ are closely related with early life gut microbiome composition and metabolic activity^10^. Thus, we hypothesized that gut microbiome functional features at baseline are associated with both POIT-induced clinical outcomes and peanut IgE levels, and that longitudinal assessment of fecal microbiomes from children in this trial would reveal mechanisms underlying variance in POIT efficacy.

## Results

### Study Population and Clinical Trial Outcomes

Details of the IMPACT trial design and outcomes have been published previously^7^. Briefly, at baseline, 146 peanut-allergic children were randomized (2:1) to either POIT or placebo treatment. After a dose escalation phase of 30-weeks, children in the POIT arm received 2,000 mg peanut protein (lightly roasted, partly defatted [12% fat]) while the placebo group received oat flour for 104 weeks (total blinded treatment period 134 weeks). Participants who passed the 5-g peanut protein, double-blind, placebo-controlled, food challenge (DBPCFC) at the end of treatment (week 134) were categorized as desensitized (D+). Independent of the DBPCFC outcome at week 134, all participants avoided peanut consumption for 24 weeks (avoidance period), and those who passed the 5-g peanut protein DBPCFC at the end of this avoidance period (week 160) were categorized as being in remission (R+). A total of 327 longitudinal fecal samples from 79 participants (Table S1) were collected at baseline (prior to POIT initiation), end of buildup (EoB), mid maintenance (MM), end of treatment (EoT), and the end of avoidance (EoA **Fig. 1A; Table S1**). Participant baseline characteristics including age, sex and study locations are reported in **Table S2**. Based on DBPCFC results at the end of treatment and end of avoidance, the IMPACT clinical trial yielded three outcome groups, Desensitized and Remission (D+R+), Desensitized and no remission (D+R-) and no desensitization and no remission (D-R-; **Fig. 1A**).

**Figure 1.**
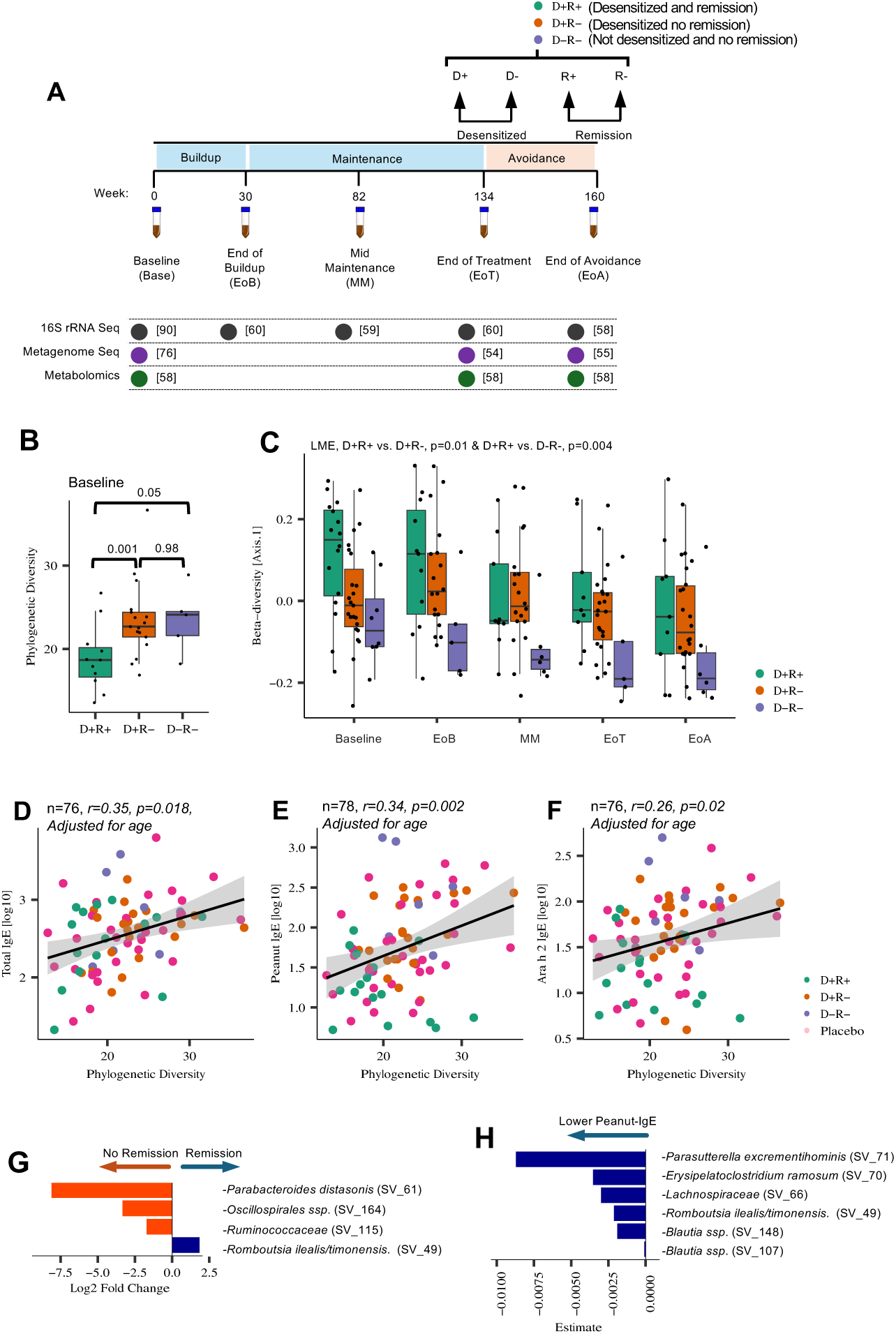
**a,** Schematic overview of IMPACT trial fecal microbiome study^9^. **b,** At baseline, prior to POIT initiation, the D+R+ group exhibit significantly lower phylogenetic diversity (α-diversity) compared to either the D+R- and D-R- groups. Wilcoxon signed-rank test, n=16 (D+R+), 23 (D+R), and 8 (D-R-). **c,** The D+R+ group exhibit a significantly distinct gut microbiota composition compared with either the D+R- or D-R- groups throughout the IMPACT trial. Linear Mixed Effect Model (*P<0.05*, not significant when adjusted for age). **d, e, f,** Baseline gut bacterial phylogenetic diversity positively correlates with baseline total IgE, peanut-specific IgE, and Ara h 2-specific IgE levels, respectively. Pearson correlation, adjusted for age at screening. **g,** Baseline differentially abundant bacterial taxa between children who achieved remission versus no remission. Generalized Mixed Model (*P.FDR<0.05,* adjusted for age). **h,** Baseline Peanut-IgE associated bacterial taxa. Generalized Mixed Model (*P.FDR<0.05*). Error bars represent standard deviation.

### Gut microbiota composition associates with peanut oral immunotherapy outcomes

The subgroup of IMPACT trial participants analyzed in our study (n=79 of 146) did not differ in age between POIT and placebo-treated groups (**Extended Data Fig. 1A**). Consistent with observations made in the parent clinical trial^7^, within the POIT-treated group, D+R+ participants were significantly younger compared to those within the two other outcome groups (D+R- and D-R-; **Extended Data Fig. 1B**). These data indicate that the subset of participants used in the current study is representative of the overall trial population. To identify potential confounding factors in our study, we initially examined relationships between variables captured in the study population and 16S rRNA sequencing-based gut microbiota profiles generated on all available longitudinally collected fecal samples (n=277 [Placebo, n=87; POIT, n=190] following quality filtering and rarefaction; see methods section for details). As expected, bacterial community α-diversity (number of taxa and their distribution) increased with advancing participant age at screening (Pearson’s correlation*, r*=0.42, *P<0.0001;* **Extended Data Fig. 1C)**. To identify other potential confounding factors, clinical and demographic variables were examined as independent terms at each time point using *PERMANOVA* based on an unweighted UniFrac distance matrix. Importantly, POIT outcomes (*P=0.008, R^2^=0.07, n=47*) and specifically remission outcome (*P=0.003, R^2^=0.04, n=47*) related to variance in gut microbiota composition only at baseline prior to POIT initiation, suggesting that gut microbiome composition prior to treatment associates with clinical outcomes. Additionally, age at screening, sample collection date, sex, and study site location significantly related to variance in gut microbiota composition at various time points throughout the trial (**Table S3**). Thus, subsequent statistical analyses performed were adjusted for these covariates (see method section for details).

### Children who develop POIT-induced remission exhibit a distinct gut microbiome throughout the course of the trial

Comparing bacterial phylogenetic diversity (α-diversity) and composition (β-diversity) over time revealed no significant difference between the POIT and placebo arms at any time point (**Extended Data Fig. 1D and 1E**), indicating that POIT does not appreciably alter fecal microbiota composition. However, within the POIT-treated group, the three distinct outcome groups exhibited significant differences in fecal microbiota α- (**Fig. 1B**) and β-diversity that were evident at baseline and sustained throughout the course of the trial (*P<0.001, R^2^=0.029, n=127*, Linear Mixed Effect; **Fig. 1C**). Specifically, the D+R+ group exhibited significantly lower α-diversity compared to D+R- and D-R- groups (*P=0.001* and *0.052*, respectively; Wilcoxon rank-sum test); this finding remained significant following adjustment for age (*P=0.043* ANOVA; **Fig. 1B and Extended Data Fig. 1F and G**). Additionally, throughout the course of the trial, participants who achieved remission (D+R+) exhibited significant differences in gut microbiota composition along the first principal component (axis 1) compared to those who did not (D+R+ vs D+R-, *P=0.03*; D+R+ vs D-R-, *P=0.003*, Linear Mixed Effect; **Fig. 1C**). This provided evidence that both gut microbiota composition and diversity are associated with POIT outcomes within the trial participants.

In the parent IMPACT trial, baseline concentrations of peanut-specific serum Immunoglobulin E (IgE) were predictive of remission^7^. We thus determined whether fecal microbiota features related to IgE levels in POIT-treated children. In age-adjusted analyses, significant positive correlations between α-diversity and serum levels of total IgE (**Fig. 1D**), peanut-specific IgE (**Fig. 1E**) and Ara h 2-specific IgE (**Fig. 1F**; *P<0.05* Pearson correlation for all) were observed, suggesting that increased fecal diversity relates to higher IgE levels that are associated with reduced likelihood of remission following POIT. Since both younger age and lower baseline peanut-specific IgE levels predicted clinical remission in the IMPACT trial, we next identified baseline bacterial Sequence Variances (SVs) associated with both peanut-specific IgE level and remission status in age-adjusted analyses. *Romboutsia ilealis/timonensis* was associated with POIT-induced remission, while *Ruminococcaceae* along with *Parabacteroides distasonis*, and *Oscillospirales* members were associated with failure to develop remission (*P.FDR<0.05*. generalized additive mix models, adjusted for age. **Fig. 1G**). *R. ilealis/timonensis* was also negatively associated with peanut-specific IgE levels at baseline (*P.FDR<0.05*. generalized additive mix models, adjusted for age. **Table S4 & Fig. 1H**) and with all component-specific Ara h-specific IgEs (Ara h 1, 2, 3 and 6 IgEs; **Extended Data Fig. 1H**). However, no significant difference in relative abundance of any SVs was observed when longitudinal analyses were performed to determine whether these findings in baseline samples were sustained through the course of the trial. Thus, 16S rRNA-based microbiota profiling provided initial evidence that gut microbiota phylogenetic diversity and composition prior to POIT initiation relate to treatment outcomes, and that the abundance of select gut microbial members associate with multiple measures of peanut allergic sensitization irrespective of participant age.

### Baseline bile acid profile associates with POIT efficacy outcomes

Distinct early-life fecal microbiome compositions associate with subsequent allergic disease development and exhibit discrete metabolic profiles that can induce allergic inflammation *in vitro*^11^. Additionally, relationships between fecal metabolite profile and food allergy have been previously described in humans^12–15^ and gut microbiomes are known to modulate host immunity through the production of metabolites, including those that can affect immunotherapy efficacy^16,17^. To determine whether the distinct fecal microbiota compositions associated with POIT outcomes exhibited divergent metabolic profiles, untargeted metabolomic analyses was performed on fecal samples collected at baseline, end of treatment, and end of avoidance from study participants (n=58 samples per visit) with sufficient available remaining sample for analysis at all three time points (**Table S1 and Fig. 1A**). Like gut microbiota composition, baseline fecal metabolite profile was associated with POIT outcome groups (*n=43, R^2^=0.07, P=0.01*), and with remission status (*n=43, R^2^=0.04, P=0.01*; *PERMANOVA,* Euclidean distance matrix, **Supplementary Table S5**).

To identify metabolites that relate to POIT outcomes, a data reduction approach, weighted gene correlation network analyses (WGCNA), was applied to identify modules of co-associated metabolites which were then related to POIT outcomes. Fifty metabolite modules (Untargeted Metabolite Modules [UMMs]; **Table S6**) were identified, 16 of which associated with POIT outcomes at baseline, end of treatment, or end of avoidance (*P<0.05* ANOVA, adjusted for age; **Fig. 2A)**. The majority of the outcome associated metabolic modules at baseline were elevated in those who did not achieve remission (**Extended Data Fig. 2A**) and included multiple lipid-containing modules, in particular bile acid and amino acid containing modules that distinguished D+R+ from D+R- and D-R- groups (**Fig. 2A**). Further supporting these observations, bile acid profiles significantly differed between POIT-outcome groups at baseline (**Fig. 2B**; *n=43, R^2^=0.10, P=0.015*, *PERMANOVA* Euclidean distance matrix), but not at the end of treatment or avoidance (**Extended Data Fig. 2B**), suggesting that the specific profile of bile acids present at the initiation of POIT influences treatment outcomes.

**Figure 2.**
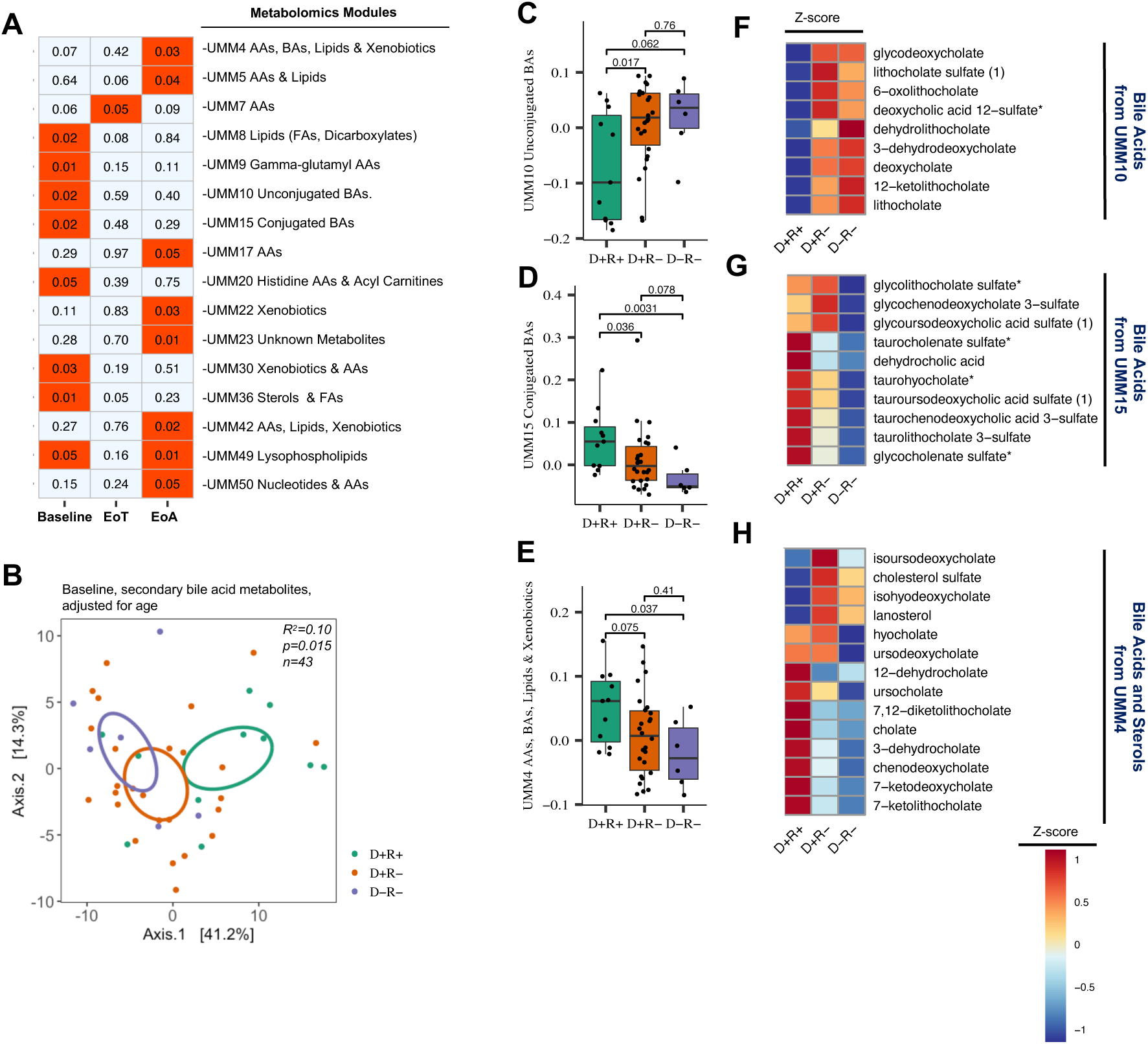
**a,** Association between untargeted metabolomics module (UMM) eigengenes and POIT outcomes (ANOVA, adjusted for age). **b,** Ordination of baseline secondary bile acid metabolites (n=43, *R*^2^ = 0.10; *P* < 0.015, adjusted for age), *PERMANOVA* analyses based on Euclidian dissimilarity metrics. **c,** Difference in baseline Module Eigengenes (ME), which were determined based on WGCNA analyses (see Method Section) and represents a measure of the joint abundance profile of a specific module, of UMM10, **d,** UMM15 Unconjugated BAs module, and **e,** UMM4 between POIT-outcome groups (Wilcoxon signed-rank test). **f,** Z-scores of each bile acid-related metabolites of UMM10, **g,** UMM15 and**, h,** UMM4 in each POIT-outcome groups (D+R+, D+R-, and D-R-). Blue colors represent low z-scores thus low abundance and red colors represent high z-score and higher abundance. Error bars represent standard deviation.

At baseline, two modules (UMM10 and UMM15) comprised primarily of unconjugated or conjugated secondary bile acids respectively, exhibited opposing relationships with POIT-induced remission (**Fig. 2C & Fig. 2D**). The module of unconjugated bile acids i.e. those lacking amino acid conjugates, was increased in those who did not achieve POIT-induced remission (**Fig. 2F**). A distinct group of secondary bile acids from UMM15 (**Fig. 2G)** and UMM4 (**Fig. 2E & Fig. 2H**) including tauroursodeoxycholic acid sulfate, glycoursodeoxycholic acid sulfate and taurolithocholate 3-sulfate were depleted in the feces of children who did not develop remission. Bile acids, produced by the liver are transformed into secondary bile acids exclusively by the gut microbiome^18^. They play crucial roles in dietary lipid absorption, regulation of lipid, glucose and xenobiotic metabolism^19^ and protect against bacterial overgrowth^18^. The secondary bile acids enriched in those who achieved remission are known to have anti-inflammatory properties and in the case of taurolithocholate, capable of down-regulating macrophage inflammatory response to antigenic stimulation^20^.

Since bile acids are drivers of gut microbiota maturation in early life^21,22^, we next investigated whether relationships existed between the bile acid modules UMM10 and UMM15 and gut microbiota features that associated with POIT outcomes. Specifically, the eigenvector (a measure of the joint abundance profile of a specific module) of UMM15, primarily comprised of bacterial-derived secondary bile acids (**Supplementary Table S6**), exhibited a significant negative relationship with fecal microbiota α-diversity (lower baseline α-diversity is associated with POIT-mediated remission; **Fig. 1B**) and a positive correlation with axis 1 of the baseline microbiota composition. In contrast the unconjugated bile acid module (UMM10) exhibited the opposite relationship, being positively correlated with fecal α-diversity and negatively correlated with axis 1 of the baseline microbiota composition (**Extended Data Fig. 2C**). These data suggest that at baseline, secondary bile acids associate with the lower gut bacterial diversity (**Fig. 1B**) and a distinct gut bacterial composition (**Fig. 1C**) that characterize children who develop POIT-induced remission.

Notably, bile acid modules that differentiated POIT outcome groups at baseline were not associated with clinical outcomes at the end of treatment or avoidance samples (**Fig. 2A**). Nonetheless, we rationalized that specific bile acids within these modules may continue to differentiate outcome groups at later time points. Leveraging generalized additive mixed models on longitudinal metabolomics samples, the UMM10 unconjugated bile acid, glycodeoxycholate was found to be in significantly higher concentrations in the feces of children for whom POIT-failed to induce remission throughout the course of the trial (*P=0.021*; **Supplementary Table S8**). Notably, glycodeoxycholate promotes innate lymphoid cell 3 secretion of IL-22^23^, which can reduce systemic absorption of peanut allergens by increasing intestinal barrier integrity, resulting in reduced antigen presenting cell encounters with peanut antigens^24^. These data indicate that significant differences in bile acid profiles exist between POIT outcome groups, particularly in baseline samples, and that glycodeoxycholate, which is known to reduce antigen exposure remains significantly elevated over time in the feces of those who fail to achieve remission following POIT treatment.

### Fecal microbiomes of non-remitting patients exhibit increased gluconeogenesis, anaerobic energy metabolism and amino acid metabolism

To investigate gut microbial pathways associated with POIT outcomes, shotgun metagenomic sequencing was performed on baseline (n=76) end of treatment, (n=54), and end of avoidance (n=55) samples (**Fig. 1A**), including all samples that had undergone parallel untargeted metabolomic analysis. Application of WGCNA to shotgun metagenomic data identified 45 shotgun metagenomic modules (SMMs; **Supplementary Table S9**), seven of which significantly associated with POIT outcomes (four at baseline and three at end of treatment (**Fig. 3A**, ANOVA, *P<0.05*, adjusted for age). At baseline, POIT-associated shotgun metagenome modules (SMMs) were primarily related to microbial growth (SMM6), energy metabolism (SMM26), peptidoglycan (SSM10), and phospholipid biosynthesis (SMM11), while at the end of treatment, long chain fatty acid production (SMM33) and gluconeogenesis and anaerobic energy metabolism (SMM43) differed across the three outcome groups (*P<0.05*, ANOVA, adjusted for age; **Fig. 3A**).

**Figure 3.**
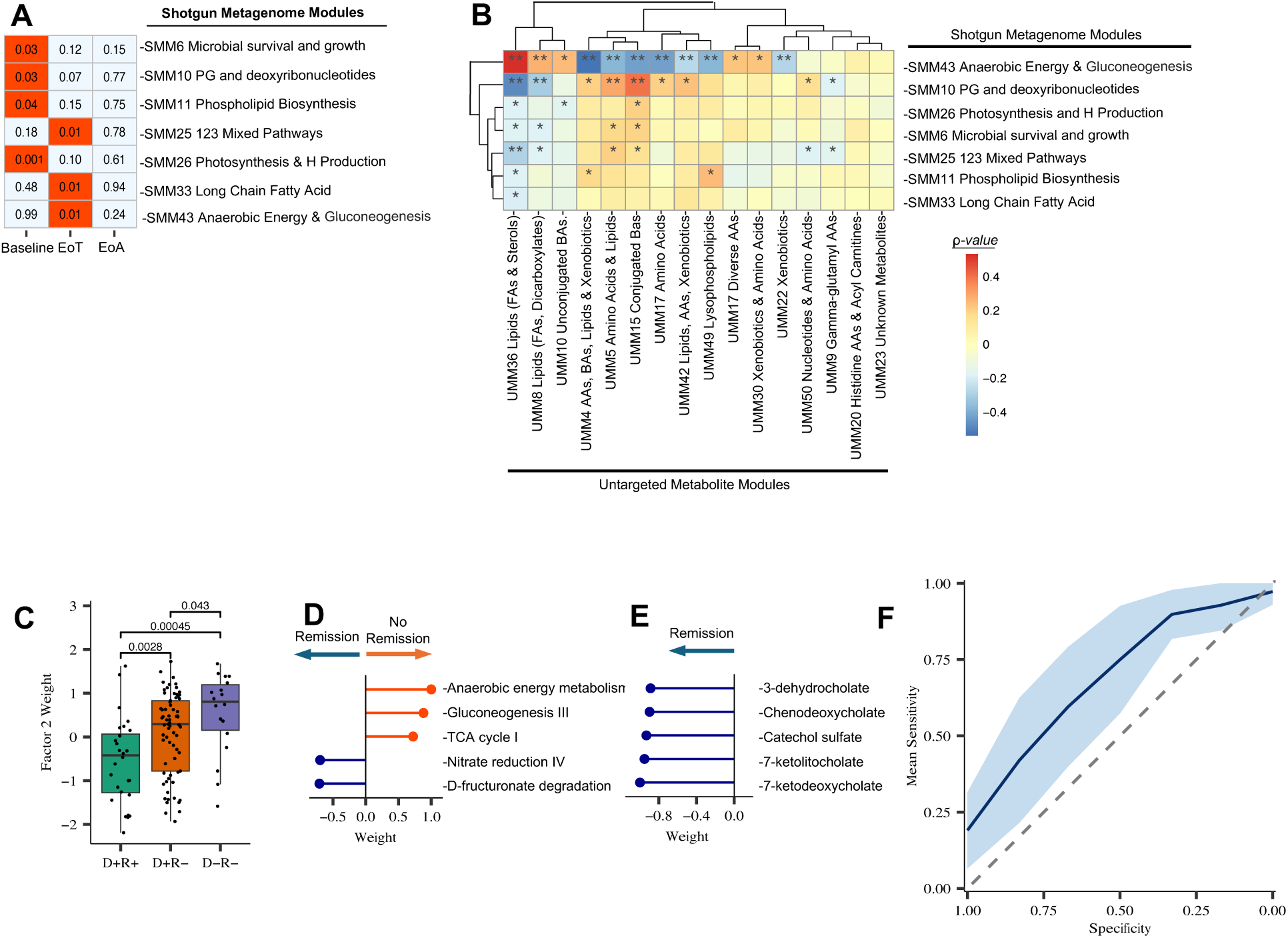
**a,** Association between shotgun metagenomic module (SMM) eigengenes and POIT outcomes (ANOVA, adjusted for age). **b,** Heatmap showing Spearman correlation results between POIT-associated Untargeted Metabolomics Modules (UMMs) and Shotgun Metagenomics Modules (SMMs). Asterisk “*” represents *P<0.05*, and double Asterisk “**” represents *P<0.01*. **c,** Factor 2 from MOFA analyses is the most significantly differential Factor between POIT response groups and weighted significantly higher in no remission groups compared to D+R+ group (*P<0.05*, Wilcoxon signed-rank test). **d,** Top 5 microbial pathways contributing the Factor 2 weight contains Gluconeogenesis and anaerobic energy metabolism pathways. **e,** Top 5 metabolites contributing the Factor 2 weight contains bile acid metabolites. **f,** The model’s predictive ability expressed as the AUC computed from 100 times repeated five-fold cross-validation. Blue line shows the average across the 100 times repeated five-fold cross-validations with the shaded area representing the 95% CI (mean AUC ± standard deviation). The dashed diagonal line represents random chance. Error bars represent standard deviation.

The gluconeogenesis and anaerobic energy metabolism module (SMM43), which was enriched in children who failed to develop POIT-induced remission at the end of avoidance (**Extended Data Fig. 2E**), was significantly correlated with 12 of the 16 metabolite modules (module eigengenes) associated with POIT outcomes (Pearson correlation, *P<0.05*; **Fig. 3B**).This included a positive correlation with the module of unconjugated bile acids (UMM10) and negative correlations with the secondary bile acid module (UMM15) and three remission-associated amino acid modules (UMM4, UMM5, and UMM43; **Fig. 3B**) that were decreased in non-responders (**Extended Data Fig. 2D)**. These data provide evidence that a fecal microbiome primarily engaged in alternate pathways of glucose production from non-carbohydrate sources and anaerobic metabolism contributes to the observed bile acid deconjugation and amino acid depletion in participants who fail to achieve POIT-induced remission.

To validate these observations, we performed a secondary, integrative data analysis on longitudinal metagenomic and paired metabolomic datasets using Multi-Omics Factor Analyses (MOFA2) which identified seven distinct factors (**Extended Data Fig. 3A**), five of which significantly differentiated POIT response groups (ANOVA, *P<0.05*; **Fig. 3C & Extended Data Fig. 3B**). Consistent with our findings, several of these factors included microbial pathways for amino acid biosynthesis that were enriched in those who achieved POIT-induced remission. One factor (Factor 2) included Gluconeogenesis and Anaerobic Energy Metabolism amongst the top five microbial pathways within this factor (**Fig. 3C, Extended Data Fig. 3C**), which were enriched in the fecal microbiome of children who did not develop POIT-induced remission (**Extended Data Fig. 3B**). In addition, secondary bile acid metabolites including 7-ketodeoxycholate, 7-ketolithocholate, chenodeoxycholate were amongst the top 5 metabolites in Factor 2, all of which were depleted in the no remission group (**Fig. 3E**). Together, these data validate that microbial gluconeogenesis and anaerobic metabolism relates with changes in the profile of secondary bile and amino acids that associate with POIT efficacy.

To assess whether the top five secondary bile acids from Factor 2, which were all increased in the remission groups at baseline, could serve as a predictive marker for POIT-induced remission, a machine learning approach with logistic regression model was employed. The baseline abundance of these five metabolites produced a moderate predictive ability (area under the curve (AUC) from 100 times repeated five-fold cross-validation, measured as mean AUC ± standard deviation (s.d.: AUC_logistic_regression_: 0.712 ± 0.081; **Fig. 3F**). To confirm our findings, a second machine learning model with random forest was employed and demonstrated similar performance (**Extended Data Fig. 3E**).

### Enhanced Microbiome Amino Acid Metabolism Associates with Failure to Induce Remission

At the end of avoidance, the majority of POIT-associated metabolite modules (five out of eight) were primarily comprised of amino acids (**Fig. 2A, Supplementary Table S6**). Several of these (UMM4, 5, 17 and 42) were decreased in children for whom POIT failed to induce desensitization and remission (D-R-; **Extended Data Fig. 2D**). Notably, amino acid profiles were significantly different among POIT outcome groups at baseline (*n=43, R^2^ = 0.08; P = 0.006;* **Fig. 4A**) and end of avoidance (*n=43, R^2^ = 0.07; P = 0.039*, **Fig. 4B**), but not at the end of treatment (*PERMANOVA* analyses, **Extended Data Fig. 3F**), suggesting that differences in dietary amino acid intake and/or microbial amino acid metabolism differentiate those who do or do not develop remission in response to POIT.

**Figure 4.**
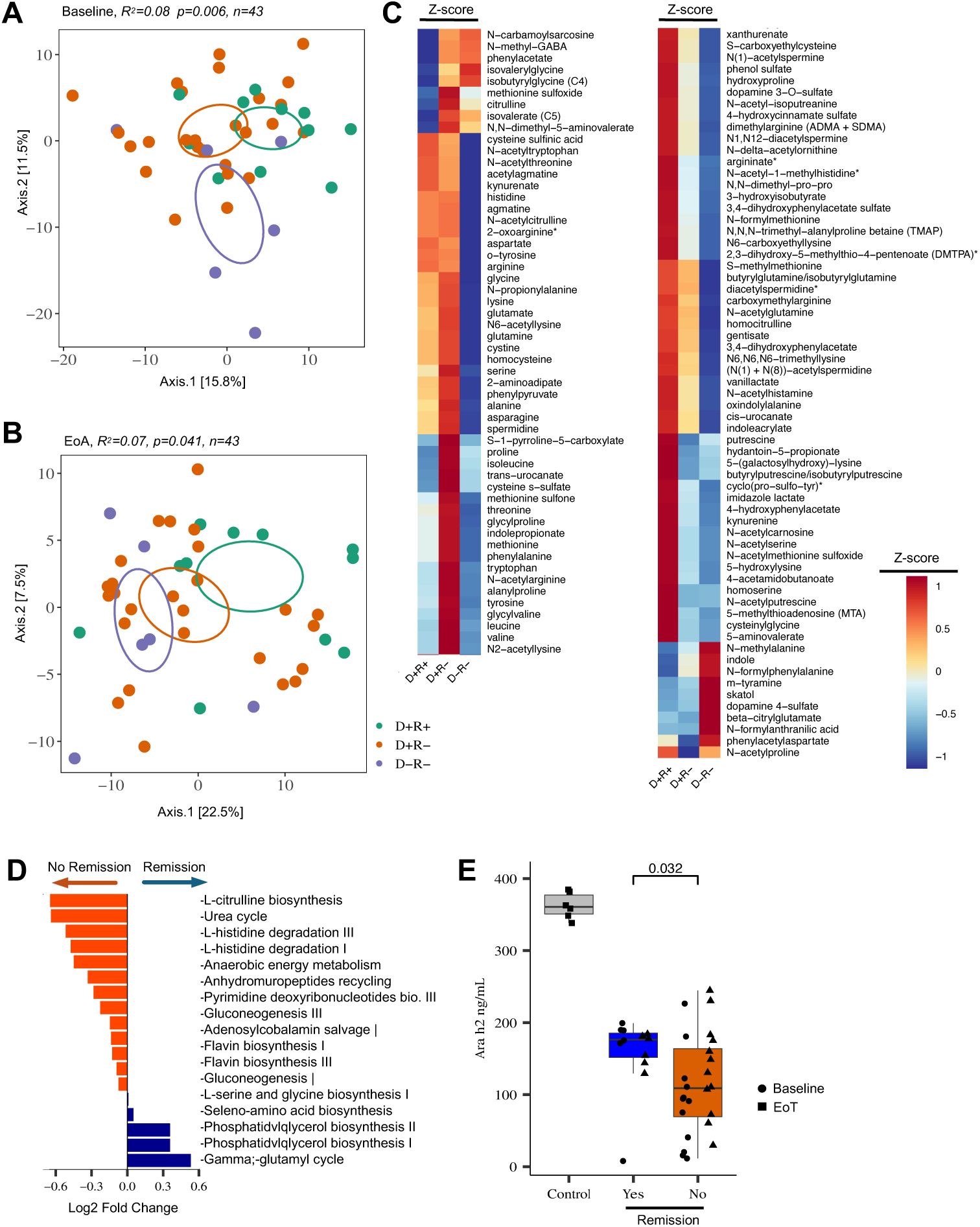
Fecal amino acid metabolite composition is distinct between POIT outcome groups at **a,** baseline (*n=43, R^2^ = 0.08; P = 0.006*), and **b,** end of avoidance (*n=43, R^2^ = 0.07; P = 0.039*). The colors representing the POIT outcome groups are as follows: green for D+R+, orange for D+R-, blue for D-R-. **c,** Z-scores of all amino acid metabolite abundance in eight modules-associated end of avoidance POIT outcome. **d,** Gut microbial pathways enriched in microbiome of children who developed remission (blue bars) versus no remission (orange bars). Generalized Mixed Models (*P<0.05, P.FDR>0.05*). **e,** Fecal microbiome of children who did not achieve POIT-induced remission have a higher capacity to metabolize peanut proteins compared to children who achieved POIT-induced remission (Wilcoxon signed-rank test). Data presented in this plot is the average of two independent experiment. Control group refers to BHI medium supplemented with peanut extract and incubated 48 h with other samples without the microbiome inoculation. Error bars represent standard deviation.

Moreover, at end of avoidance, POIT response-associated metabolite modules contained a total of 117 amino acids and their derivatives, 68 of these belonged to UMM4 (**Table S6**), which was significantly reduced in the D-R- group (**Extended Data Fig. 2D**). The majority of these amino acid metabolites’ abundances were decreased in relative concentration in the feces of children who failed to achieve remission (**Fig. 4C**) Notably, increased concentrations of microbial-derived branched-chain amino acid fermentation end products such as skatol and indole^25^ were evident in both the D+R- and D-R- groups (**Fig. 4C**) indicating that the observed reduction in amino acid concentrations is due to increased microbial amino acid utilization capacity in those who failed to achieve POIT-induced remission.

Amino acids represent a major energy source for anaerobic gut bacteria^26^, and select microbes are capable of harvesting amino acids by deconjugating primary bile acids^27^. Decreased fecal amino acid concentrations and increased anaerobic energy and gluconeogenesis metabolism in POIT-treated children who failed to achieve remission prompted us to investigate whether their gut microbiomes encoded a distinct or enhanced capacity for amino acid utilization. Using generalized additive mixed models on longitudinal microbial pathway abundance data, we found that the microbial pathways associated with amino acid metabolism including L-histidine degradation, anerobic energy metabolism, L-citrulline biosynthesis (Arginine degradation), and gluconeogenesis (which uses non carbohydrate sources, including amino acids for energy production^28^) were enriched in the gut microbiomes of children who did not achieve remission. In contrast, and consistent with metabolite modules associated with remission, D+R+ children possessed fecal microbiota that encoded pathways involved in amino acid biosynthesis (generalized additive mix model, *P<0.05*, but *P.FDR>0.05*; **Fig. 4D & Table S8**). These data indicate that gut microbiomes with enhanced capacity for gluconeogenesis and amino acid metabolism, result in depletion of immunomodulatory amino and secondary bile acids that associate with POIT failure.

Because peanut antigen exposure is critical to develop immunological tolerance to peanut^3,29^ and children who failed to achieve remission exhibited a gut microbiome with enhanced capacity for amino acid metabolism (**Fig. 4D**), we hypothesized that their gut microbiome may also have increased capacity for peanut metabolism, effectively reducing available antigen. To test this, stabilized *in vitro* fecal microbiome cultures from participants in each of the outcome groups (D+R+, n=12, D+R-, n=12, D-R-, n=12) were developed as previously described*^30^* and co-incubated with peanut extract under anaerobic conditions prior to Ara h 2 quantification by ELISA. Fecal microbiomes of all participants, regardless of remission outcome, exhibited the capacity to metabolize Ara h 2 peptides, one of the most proteolysis-resistant peanut protein antigens^31^. However, the microbiome of those who failed to achieve remission exhibited a significantly increased capacity to metabolize Ara h 2 peptides compared to those who achieved remission, indicating that enhanced microbial metabolism of allergenic peanut peptides associates with POIT failure (**Fig 4E**).

## Discussion

Integrated analyses of 16S rRNA data generated from fecal samples longitudinally collected from IMPACT participants (Placebo, n=87; POIT, n=190) provide evidence that microbial activities in the fecal microbiome prior to POIT initiation relate to treatment outcomes. The data also indicate that the composition of the gut microbiome is distinct over a three-year treatment period in those who do or do not experience peanut allergy remission following POIT, suggesting that microbial-mediated changes in immune function are associated with distinct trajectories of microbiome development and POIT outcomes in study participants. Functional analyses of fecal microbiomes indicate that bile acids, specifically secondary bile acids enriched in baseline samples, associate with POIT-induced remission. Microbial-derived secondary bile acids serve as hormones that regulate cholesterol metabolism and influence energy balance via nuclear and G-protein-coupled receptors^32,33^ that also shape innate immune response^34,35^. Studies have demonstrated that the bile acid pool regulates colonic FOXP3+ regulatory T (Treg) cells that express the transcription factor RORγ^36^. Bile acids found to be increased in baseline samples of participants who experienced peanut allergy remission in our study are known to have allergy protective effects; for example, tauroursodeoxycholic acid attenuates allergic inflammation by inhibiting unfolded protein response transducers^37^.

Children for whom POIT failed to induce remission exhibit a significantly distinct gut microbiome, enhanced for gluconeogenesis and anaerobic metabolism, indicating that these gut microbiomes derive energy from alternative non-carbohydrate substrates. Our data indicate that the gut microbiome of these children is enhanced for amino acid metabolism. Evidence for this comes from the increased pools of deconjugated (primary) bile acids and a significant depletion of secondary bile and amino acids in the feces of these children. Primary bile acids are most commonly conjugated to the amino acids glycine and taurine to produce glycocholic and taurocholic acids respectively^18^, which are subsequently converted to immunoregulatory secondary bile acids by colonic bacteria^18^. Primary bile acids, specifically chenodeoxycholic acid, was recently shown to promote food sensitization via activation of a retinoic acid response element in dendritic cells, to promote food allergen specific IgE and IgG1^38^. Enhanced colonic microbial capacity to harvest amino acids from conjugated bile acids results in increased concentrations of the deconjugated forms, essentially reverting them back to their primary bile acid state. Thus, our findings indicate that a gut microbiome that primarily derives energy from amino acid fermentation results in the depletion of immunomodulatory amino and secondary bile acids both of which associate with POIT remission failure.

It is well established that peanut allergen exposure is critical to build tolerance and prevent allergy development^3^. Our data suggests that a second microbial-derived mechanism of POIT failure appears to involve reduced peanut antigen exposure. Food allergies and intolerances are typically driven by specific protein motifs in foods such as Ara h peptides in peanuts, casein and beta-lactoglobulin in cow’s milk and tropomyosin proteins in shellfish^39^. Ingested allergens typically undergo enzymatic breakdown in the oral cavity, stomach, and small intestine^40^ prior to interacting with antigen-presenting cells^41^. However, certain key antigenic peanut peptides e.g. Ara h 2 peptides, are highly resistant to proteolysis^42^, making it likely that they survive transit through the upper gastrointestinal tract to the distal colon. The extent of peanut protein digestion determines the concentrations and profile of antigenic peptides available for presentation by antigen presenting cells. Our data indicates clear differences in amino acid metabolism capacity and peanut degradation in participants who did or did not achieve remission suggesting that differences in distal gut microbial protein catabolism may impact the quantity and profile of antigenic peanut peptides available to promote tolerance development.

Gut microbial protein metabolism has never been explored in the context of food allergy, particularly related to treatment-induced efficacy outcomes. Elucidating the impact of gut microorganisms on allergic food proteins may pave the way to develop more effective immunotherapeutic approaches either by targeting gut microbiome functions or by protecting immunotherapeutic peanut proteins from microbial metabolism by encapsulating them in food-grade colloidal systems. A similar encapsulation system for gluten immunotherapy is currently being tested in several clinical trials^43,44^, which so far have demonstrated safety and efficacy. Our study highlights the potential role of the gut microbiome in POIT efficacy outcomes and suggests that it could serve as both a prognostic biomarker to identify those for whom POIT may be most successful and as a therapeutic target to improve rates of POIT-induced remission.

## Materials and Methods

### Clinical trial description and study population

Full details of the IMPACT clinical trial (NCT01867671) have been previously described^45^.

### Sample collection, DNA extraction, 16S rRNA library preparation and sequencing

Stool samples were collected from participants at home and stored at clinical collection sites at -80 °C. Three hundred twenty-seven samples (n=327) were shipped to the University of California San Francisco (UCSF), on dry ice, where they were also stored at -80 °C until processed. Investigators of this study were blinded and did not have access to the metadata until initial 16S rRNA data generation. Thus, all 327 samples were included for 16S rRNA sequencing. Three hundred two (302) samples generated 16S rRNA data since some samples failed PCR amplification or did not pass quality filtering and rarefaction during 16S rRNA analyses. Two hundred seventy-seven (277) out of 302 samples belonged to participants that completed the POIT trial until the end of the avoidance, and therefore, they were used in 16S rRNA data analyses.

DNA was extracted from all stool samples using a modified cetyltrimethylammonium bromide (CTAB) buffer extraction protocol as previously described^11,46^. The variable region 4 (V4) of the 16S rRNA gene was amplified using 1 ng μl^−1^ of DNA template using 515F and 806R primer pairs as previously described^47^. Amplicon concentrations were normalized using SequalPrep™ Normalization Plate Kit (Thermofisher Scientific), quantified using the Qubit 2.0 Fluorometer and the dsDNA HS Assay Kit (Life Technologies) and pooled at 5 ng per sample which was purified using AMPure SPRI beads (Beckman Coulter). 2 nM of library was spiked with 30% of PhiX control v3 (Illumina). The denatured libraries and PhiX were diluted to 20 pM, and 1.5 pM were loaded onto the Illumina NextSeq 500/550 v2.5 High Output cartridge.

Sequence data was processed as previously described^48^. Forward and reverse reads were demultiplexed by using Quantitative Insights Into Microbial Ecology (QIIME 1.9.1)^49^. Samples sequences with more than two bases having a Q-score less than 30 were truncated. As recommended by the Divisive Amplicon Denoising Algorithm 2 (DADA2) v1.16 protocol in R with the following modifications: Reads were maintained if they exhibited a maximum expected error of two and a read length of at least 150 base pair (bp) using the *filterAndTrim* function in the *dada2* package^50^. Reads were dereplicated and errors were learned on 1 × 10 reads, from samples chosen at random. Finally, chimeras were identified using the ‘‘consensus’’ method. Paired reads were merged with a minimum overlap of 25 bp, and reads were aggregated into a count table. Any V4 sequences abnormally short or long (±5 bp from the most frequently observed bp length; here: 253 bp) were also removed. We assigned taxonomic classifications to Sequence Variants (SVs) using *assignTaxonomy* in the *dada2* package and an 80% bootstrap cutoff, utilizing the SILVA v132 database^51^, and species identification with *assignSpecies* at 100% identity. All species achieving an exact match were recorded, and the first in the list was used for descriptive purposes. Once these steps were completed for each run, all runs were combined into a complete SV table. A phylogenetic tree was constructed using *phangorn^52,53^* and *DECIPHER* packages^54^. The SV table was then filtered only to variants belonging to the kingdom Bacteria. Variants were also removed if they were present in less than 0.001% of the total number of observed sequences reads. Next, we employed methods to remove potential contaminants based on SVs present in negative controls. Specifically, SVs were removed if they were present in greater than 15% of the negative controls and less than 15% of the samples^48^ (primarily *Pseudomonas* SVs). For the remaining sequence variants in negative controls, the mean of the read count for each was calculated, rounded upward to the nearest whole number and subtracted for each of these SVs in the dataset. Any remaining negative control SVs were subtracted from samples using the maximum read count across negative controls. Data was representatively rarefied at 35,000 reads per sample, a level selected to optimize sample count and community coverage.

### Metagenomic processing and data analysis

One-hundred eighty-five samples (n=76 Baseline, n=54 EoT, n=55 EoA, **Fig. 1A)**, were chosen among the DNA samples extracted for 16S rRNA sequencing including samples that went through untargeted metabolomic analyses. Fecal samples selected had sufficient remaining material for paired metagenomic and metabolomic profiling. Extracted DNA was sent to the Omega Bioservices Sequencing Laboratory (Norcross, GA, USA) for shotgun metagenome sequencing. DNA concentration was measured using the QuantiFluor dsDNA System on a Quantus Fluorometer (Promega, Madison, WI, USA). A Kapa Biosystems HyperPlus kit (Kapa Biosystems, Wilmington, MA, USA) was used for library construction. Briefly, 50 ng of genomic DNA was enzymatically sheared according to the manufacturer’s instructions. DNA fragment ends were repaired, 3’ adenylated, and ligated to adapters. The resulting adapter-ligated libraries were PCR-amplified. PCR product was cleaned up from the reaction mix with magnetic beads. Then, Illumina libraries were quantified using the Qubit 2.0 Fluorometer with the dsDNA High Sensitivity Assay Kit (Life Technologies, Grand Island, NY) and pooled at equal molar concentrations. The final pooled libraries were submitted to the Center for Advanced Technology (CAT) at the University of California San Francisco. The pooled libraries were sequenced using the Illumina NovaSeq 6000 in a 2×150 bp paired-end run protocol targeting minimum 60,000,000 raw reads per sample in total.

Raw sequences from all lanes were merged into a concatenated file for each sample. Raw FASTQ files underwent FASTQC^55^ and quality and contaminant filtering using *bbTools* v38.73. Specifically, *bbduk* trimmed Illumina adapters, removed any PhiX contamination, filtered low-quality sequences, and employed trimming after a Q score less than 15 from both the 3′ and 5′ directions. Finally, *bbmap* removed reads mapping to the human genome using GRCh38^56^ as the reference database. The median number of raw reads per sample was 97,502,238 (IQR 30,132,152). The median number of reads following Q15 quality trimming and filtering human DNA using Bbduk v38.73 (https://sourceforge.net/projects/bbmap/) was 13,367,212 (IQR 2,073,330). All analyses were performed on quality-filtered reads. HUMAnN 3.0 pipeline was used to identify genes^57^, level4ECs and functional MetaCyc pathways from the short-reads, and to normalize outputs into copies per million (CPM).

### Untargeted Metabolomics Analyses

Among the samples that went through shotgun-metagenome analyses, 174 (n=58 Baseline, n=58 EoT, n=58 EoA, **Fig. 1A**) matching samples were available for untargeted metabolomics analyses. Two hundred milligrams of stool per sample was submitted to Metabolon Inc. (Durham, NC) for ultrahigh performance liquid chromatography/tandem mass spectrometry (UPLC–MS/MS) and gas chromatography–mass spectrometry (GC–MS) using their standard protocol (http://www. metabolon.com/). Briefly, samples were homogenized and subjected to methanol extraction then split into aliquots for analysis by ultrahigh performance liquid chromatography/mass spectrometry (UHPLC/MS) in the positive (two methods) and negative (two methods) mode. Metabolites were then identified by automated comparison of ion features to a reference library of chemical standards followed by visual inspection for quality control (as previously described^58^. Compounds were compared to Metabolon’s in-house library of purified standards, which includes more than 3,300 commercially available compounds. For statistical analyses and data display, any missing values are assumed to be below the limits of detection; these values were imputed with the compound minimum (minimum value imputation). For network and statistical analyses, normalized, imputed, and log transformed area under curve dataset was used.

### *In vitro* Fecal Microbiome Metabolism of Peanut

Stool samples from IMPACT participants were prepared for culture as described previously*^30^*. Briefly, stool samples from 36 patients (D+R+, n=12, D+R-, n=12, D-R-, n=12) with sufficient paired baseline and end of treatment material for the experiment were thawed on ice. All fecal processing was completed under aerobic conditions. Stools were resuspended in Brain Heart Infusion (BHI) media at a ratio of 10 ml/g stool prior to vigorous vertexing for 5 min and filtering with a 50 µm cell strainer and storage at -80°C following 25% (volume/volume) glycerol addition. A total of 10 µL of prepared feces was used to inoculate 1 mL of BHI medium supplemented with 8 µL peanut extract (1/10 weight/volume in 50% glycerin, Hollister-Stier) and incubated for 48 hours at 37°C under anaerobic conditions. Following 48 hours incubation, microbiome cultures were centrifuged at 3,200 *g* for 10 min and filtered through 0.22 µm filters. Ara h 2 peptide concentrations were determined using an Enzyme-Linked Immunosorbent Assay (ELISA) according to manufacturer instructions (Indoor Biotechnologies, Charlottesville, VA).

### Statistical analyses

Statistical analyses were performed in the R statistical programming language version 4.3.2. Phylogenetic diversity (Alpha diversity) was calculated in QIIME and was expressed as Faith’s phylogenic diversity metric, using the vegan and picante packages in R. Wilcoxon tests were calculated in R. For beta-diversity (microbiome composition), distance matrices based on unweighted UniFrac for 16S rDNA data and Euclidian for metabolomics dataset were generated using the *distance* function from *phyloseq* v1.30.0^59^ and ordinated into two-dimensional space using the *pcoa* function from the *ape* v5.3 package*^60^*. Permutational Analysis of Variance tests (*PERMANOVA*; *R^2^* and *P* values) were generated for independent terms with 1000 permutations using *adonis2* from the *vegan* package v2.5-6^61^. Pearson correlations and *P* values were calculated and corrected for potential confounding factors such as age at screening, using the cor.test function in R. When samples were used from multiple time points, for example, in Linear Mix Effect (LME) models on longitudinal samples, only age was adjusted, and stated in figure legends.

### Generalized Additive Mix Model

Generalized Linear Mix Models were used on longitudinal microbiome data to determine differences in microbial taxa, microbial pathways, metabolites between POIT outcome groups (D+R+, D+R-, D-R-) and remission outcome (Yes or No), using Linear Model, Compound Poisson Linear Model, Poisson, Negative Binomial, Zero-Inflated Negative Binomial, and Tweedie models depending on data distribution. False-discovery corrections were made using the Benjamini-Hochberg method.

### Weighted Gene Correlation Network Analyses

Co–occurrence networks of microbial pathways and metabolites were constructed using weighted correlation network analysis (WGCNA) with the R package *WGCNA^62^* to find modules of highly interconnected, mutually exclusive metabolites. Pearson correlations were used to determine inter–metabolite and inter–microbial pathway relationships, where modules are composed of positively correlated metabolites. We constructed a signed network using specific parameters (power = 7, reassignThreshold = 0, mergeCutHeight = 0.25), by applying hierarchical clustering and topology overlap measures (TOM). The minimum module size was set to five for metabolomics and one for metagenomics data. Module eigengenes (MEs) were defined as the first principal component of a given module and considered as a representative measure of the joint abundance profile of that module. Each module eigengenes was used to test the association between its respective module and POIT-outcomes using ANOVA. Module membership was used to determine the interconnectedness of each metabolite or microbial pathways to its assigned module and to identify “hub” metabolite or microbial pathways: this was defined as the correlation between each metabolite or microbial pathways and the Module eigengenes (MEs) (strong positive values indicate high interconnectedness) as previously described^9^.

### Multiomics Factor Analyses (MOFA2)

MOFA uses multi-omics data from the same set of samples as input and generates a model that infers a set of “Factors” that best explain patterns of covariation across samples^63^. Details of methodology can be found in the original publication^64^. As input for the MOFA model, we used untargeted metabolomics (1538 metabolites) and shotgun metagenomics datasets (518 features). All inputs were normalized by centralized log normalization. When fitting the model, we selected for the top factors ordered by the mean fractional variance explained across omic modalities (that is, factor 1 contributed the most, and factor 7 contributed the least to mean fractional variation; **Extended Data Fig. 3A**). When testing factor values for statistically significant differences between POIT outcome groups we used a two-tailed Mann–Whitney *U*-test. **Extended Data Fig. 3B)**. Top five features of metagenome and metabolome datasets from significant factors were displayed (**Extended Data Fig. 3C**).

### Machine Learning Model with Logistic Regression and Random Forest

For the predictive metabolite analysis, normalized abundance of top five metabolites from Factor 2 of the MOFA2 analyses were processed with the mikropml R package (https://CRAN.R-project.org/package=mikropml)^65^. We used Random Forest (rf) and Logistic Regression functions (glmnet) with Remission (Yes versus No) as an outcome using 50% of the samples as training set and 50% as the test set. Model performances were evaluated with repeated k-fold cross-validation (tenfold, 10 repetitions) and parameters were tuned by choosing mtry and values between 1 and the square root of the total number of variables. Model training was accomplished with the caret R package (https://topepo.github.io/caret/), mtry and lambda values that determined the highest model accuracy were chosen as input to Random Forest and Logistic Regression analysis, respectively. Variable importance was assessed with permutations (100 iterations). Full results are reported in **Supplementary Table S10**.

## Data Availability

All data produced in the present study are available upon reasonable request to the authors

## Availability of data and materials

The sequencing data generated from untargeted metabolomics, shotgun metagenomes and amplicon sequencing for this study will be deposited to the NCBI SRA database. Additional data can be shared upon request.

## Acknowledgements

Research reported in this publication was supported by the National Institute of Allergy and Infectious Diseases of the National Institutes of Health under Award Number UM1AI109565 and by the S.V.L’s research program which is funded by AI128482, AI148104, UM1AI160040 and AI089473. The content is solely the responsibility of the authors and does not necessarily represent the official views of the National Institutes of Health. M.Ö. is supported by postdoctoral T32 fellowship 2T32DK007762-46. We thank Rebecca L. Knoll, and Elad Deiss-Yehiely for their internal review of this article. We also thank Immune Tolerance Network (ITN) Leadership, IMPACT study team, and study participants for making this study possible.

## Author contributions

Conceptualization, M.Ö. and S.V.L.; methodology, M.Ö., D.L.L., C.L.G., A.L., and S.V.L.; data analysis, M.Ö.; investigation, M.Ö. D.L.L., C.L.G., A.L., L.M.W., C.B., S.S., S.M.J., and S.V.L writing and editing original draft, M.Ö. and S.V.L.; writing review M.Ö., C.L.G., and S.V.L.; data visualization, M.Ö., supervision, S.V.L., funding acquisition, S.V.L.

## Ethics Statement

This study was approved by the Office of Human Research Ethics (OHRE), University of North Carolina, Chapel Hill on April 9, 2013. The parent study titled, “IMPACT: Oral Immunotherapy (OIT) for Induction of Tolerance and Desensitization in Peanut-Allergic was a randomized, double-blind, placebo-controlled, multi-center study comparing peanut oral immunotherapy (OIT) to placebo. Informed consent was obtained from a parent or guardian of all participants.

## Extended Data

**Extended Data Figure 1.**
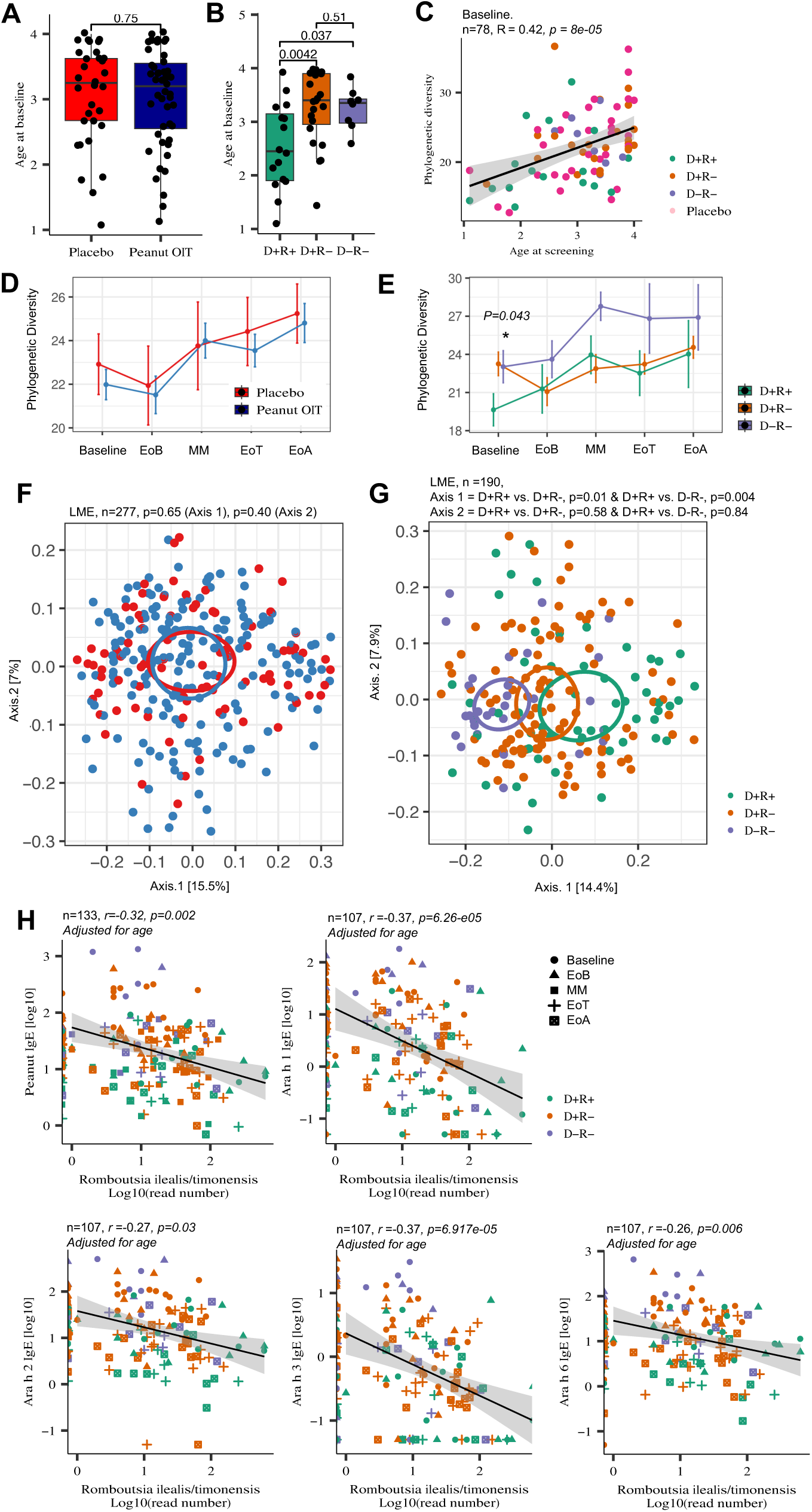
**a,** There was no significant difference in age between Placebo and POIT groups at baseline (ANOVA, P>0.05). **b,** Participants who achieved POIT-induced remission in the current study were significantly younger compared to participants in D+R- and D-R- groups, similar to the original study (ANOVA*, P<0.05)*. **c,** Phylogenetic diversity positively correlates with participant age. (*P<0.05*; Pearson correlation). **d,** Phylogenetic diversity *was* similar between Placebo and POIT arms throughout the IMPACT trial *(Linear Mix Model, adjusted for age)*. **e,** D+R+ has a lower bacterial phylogenetic diversity at baseline compared to D+R- and D-R- groups *(P=0.043, Linear Mix Model, adjusted for age)***. f,** Gut bacterial composition is similar between Placebo and POIT groups throughout the IMPACT trial (LME n=277; Placebo=87, POIT=190). **g,** Gut bacterial composition is distinct between POIT outcome groups (n=190; D+R+ =54, D+R- =106, and D-R- =30; LME). **h,** Abundance of *Romboutsia ilealis* negatively correlates with all measured peanut-specific and component specific IgE levels; Ara h 1, Arah 2, Ara h 3, and Ara h 6 (Pearson correlation, adjusted for age, *P<0.05*). LME: Linear Mix Effect. Error bars represent standard deviation.

**Extended Data Figure 2.**
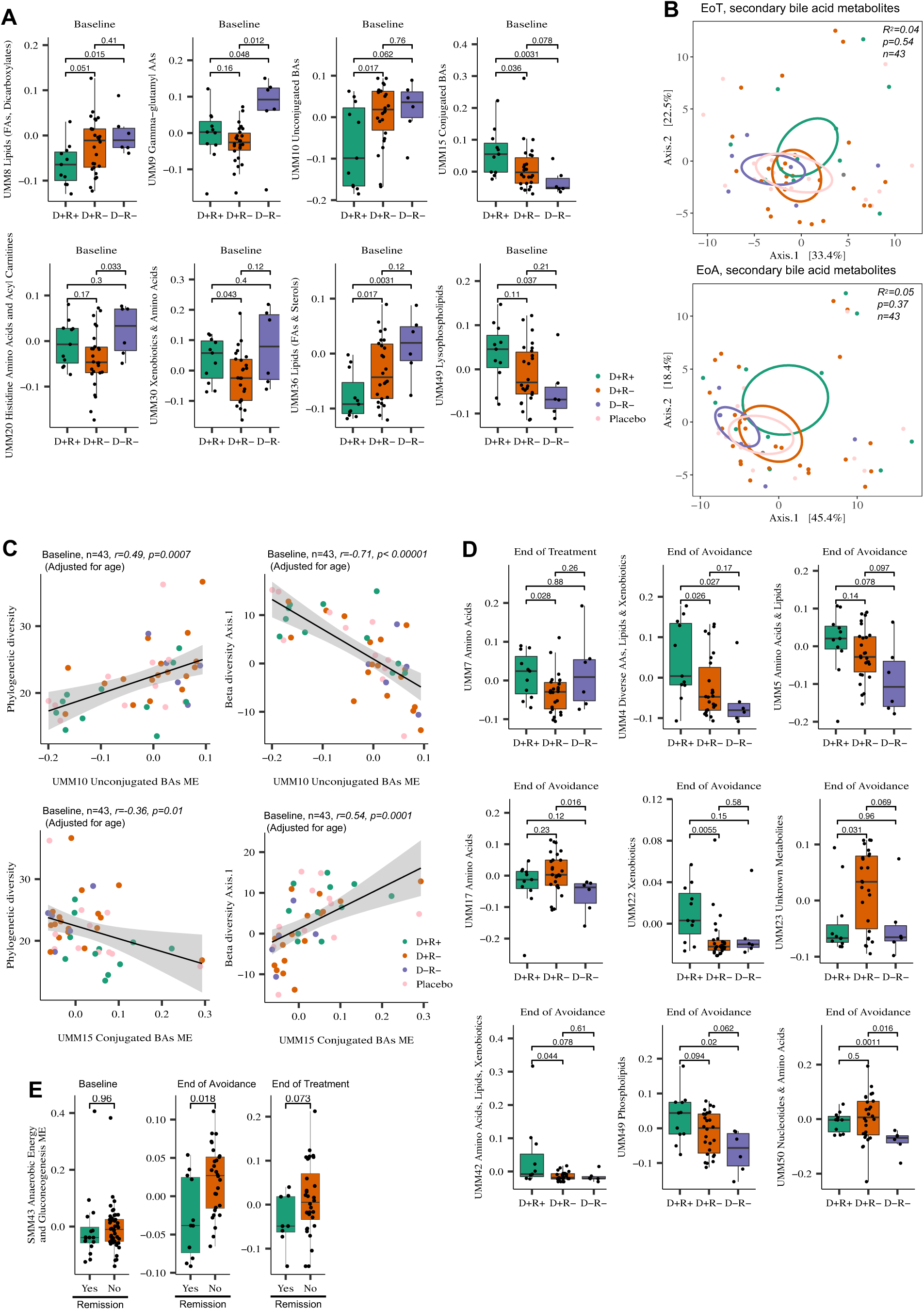
**a,** Difference in baseline module eigengenes (ME) of POIT outcome-associated untargeted metabolomics modules (Wilcoxon signed-rank test). **b,** Fecal bile acid metabolite composition is not different between POIT-outcome groups at the end of treatment and avoidance. Ordination of end of treatment and avoidance secondary bile acid metabolites, *PERMANOVA* analyses based on Euclidian dissimilarity metrics (P> 0.05). **c,** The bile acid modules (UMM10 and UMM15), which are significantly associated with POIT-outcome, correlate with phylogenetic diversity and gut microbiome composition (*P<0.05*, Pearson Correlation, adjusted for age). **d,** Difference in end of treatment and avoidance module eigengenes (ME) of POIT outcome-associated untargeted metabolomics modules (Wilcoxon signed-rank test). Error bars represent standard deviation.

**Extended Data Figure 3.**
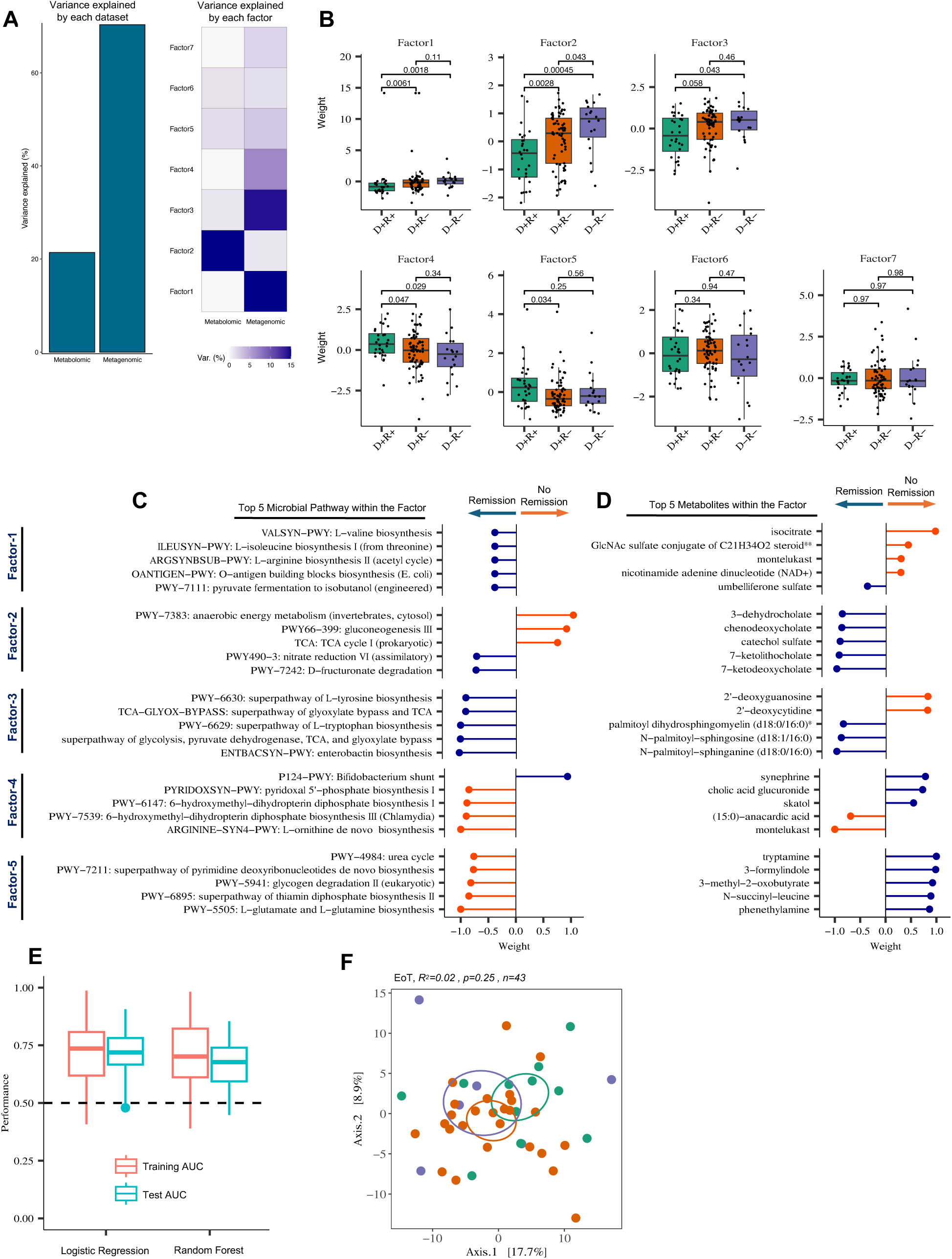
**a,** MOFA2 analyses, variance explained by each omics datasets; metabolomics and metagenomics. **b,** Seven MOFA2 factors were identified, five of which (Factors 1-5) were significantly different between POIT outcome groups (*P<0.05*, (Wilcoxon signed-rank test). **c,** Top five microbial pathways contributing to weight of each factor. **d,** Top five metabolites contributing to weight of each factor. **e,** Comparison of average AUC between logistic regression and random forest models in predicting remission outcome based on five metabolites from Factor 2. **f,** Fecal metabolite composition is not different between POIT outcome groups at the end of avoidance (Euclidian distance matrix. *n=43, R^2^ = 0.02; P = 0.25*). Orange and blue lollipop colors represent negative and positive effect on factor weight, respectively. Error bars represent standard deviation.

